# Assessment and optimization of respiratory syncytial virus prophylaxis regimens in Connecticut, 1996-2013

**DOI:** 10.1101/2020.07.13.20152215

**Authors:** Ben Artin, Virginia E. Pitzer, Daniel Weinberger

## Abstract

**Background:** Respiratory syncytial virus (RSV) causes seasonal respiratory infection, with hospitalization rates of up to 50% in high-risk infants. Palivizumab provides safe and effective, yet costly, immunoprophylaxis. The American Academy of Pediatrics (AAP) recommends palivizumab only for high-risk infants and only during the RSV season. Outside of Florida, the current guidelines do not recommend regional adjustments to the timing of the immunoprophylaxis regimen. We investigate the benefits of adjusting the RSV prophylaxis regimen in Connecticut based on spatial variation in the timing of RSV incidence.

**Methods:** We obtained weekly RSV-associated hospital admissions by ZIP-code in Connecticut between July 1996 and June 2013. We estimated the fraction of all RSV cases in Connecticut occurring during the period of protection offered by RSV immunoprophylaxis (“preventable fraction”) under the AAP guidelines. We then used the same model to estimate protection conferred by immunoprophylaxis regimens with alternate start dates, but unchanged duration.

**Results:** The fraction of RSV hospitalizations preventable by the AAP guidelines varies by county because of variations in epidemic timing. Prophylaxis regimens adjusted for state- or county-level variation in the timing of RSV seasons are modestly superior to the AAP-recommended regimen. The best alternative strategy yielded a preventable fraction of 95.07% (95% CI: 94.69 – 95.44%), compared to 94.07% (95% CI: 93.65 – 94.46%) for the AAP recommendation.

**Conclusion:** Initiating RSV prophylaxis based on state-level data may improve protection compared with the standard AAP recommendations. In Connecticut, county-level recommendations would provide only a modest additional benefit while adding complexity.

## 1 Introduction

Respiratory syncytial virus (RSV) causes upper respiratory tract infections in people of all ages. In adults, RSV infection is typically asymptomatic or gives rise to mild respiratory illness. However, RSV infection often incites more serious respiratory illness in children, with hospitalization rates as high as 4.4% in infants with no comorbidities, making RSV a leading cause of hospitalization among infants.^1–4^

For infants, risk factors of serious illness and hospitalization due to RSV infection include prematurity, chronic lung disease of prematurity, congenital heart disease, anatomic pulmonary abnormalities, neuromuscular disorders, trisomy 21, and immunocompromised status.^5^ The hospitalization rate due to RSV among high-risk infants is anywhere from two to ten times higher than among infants with no comorbidities.^4^

A monoclonal antibody (palivizumab) is available in the US for prevention of RSV infection in high-risk infants less than 2 years old. Palivizumab is administered via intramuscular injection and requires five monthly doses, priced at $1500-$3000 per dose.^6,7^ Palivizumab has been found to be effective in reducing hospitalizations due to RSV; in double-blinded trials in high-risk infants, it reduced the hospitalization rate by 45-55% compared to placebo.^8,9^ It has also been found to be safe and well-tolerated.^8^ Owing to the high cost, the 2014 revision of the American Academy of Pediatrics (AAP) RSV prophylaxis guidelines recommends palivizumab only for high-risk infants and only during the RSV season.^5,10,11^

RSV seasonality and spatial variability have been established throughout the United States, based on the CDC’s National Respiratory and Enteric Virus Surveillance System data; RSV has been found to have an annual or biennial cycle with regionally variable seasonal onset ranging from late August to mid-December and offset ranging from mid-March to late April.^12–14^ In Connecticut, previous analysis of RSV-associated hospital admissions has shown spatial variability in the timing of peak RSV incidence, with epidemics occurring earlier in large urban areas and later in sparsely-populated rural areas.^15^ The AAP acknowledges the potential significance of spatial and temporal variation in RSV incidence, but — outside of Florida, whose RSV season leads most of the rest of the nation by approximately two months — does not recommend local variance from nationwide prophylaxis guidelines.^5^

Motivated by the AAP’s recognition of the significance of spatial variation in formulating prophylaxis guidelines and the known spatial variability of RSV incidence in Connecticut, we aim to determine whether protection due to RSV prophylaxis would be enhanced by having localized guidelines for the initiation of prophylaxis by addressing the following questions:

- To what extent does county-level variability of RSV incidence in Connecticut impact the effectiveness of the AAP-recommended prophylaxis regimen?
- Would county- or state-specific alternative regimens increase the benefits of prophylaxis in comparison to the AAP-recommended regimen without increasing the total cost of the regimen?
- Is there important year-to-year variation in timing of RSV season in Connecticut?

## 2 Methods

### Source of data

We obtained data consisting of all hospitalizations among children <2 years old in Connecticut from 1996 to 2013 from the Connecticut State Inpatient Database through the Connecticut Department of Public Health (CT DPH). The relevant variables in the data were ZIP code of residence, patient age, ICD-9 defined diagnoses (up to 10 diagnoses per hospital admission), and week and year of hospital admission. The study was approved by the Human Investigation Committees at Yale University and the CT DPH. The authors assume full responsibility for analyses and interpretation of the data obtained from CT DPH.

### Definitions

We defined an RSV-associated hospitalization in Connecticut to be a hospital stay for which:

- the postal code of residence listed the in patient’s medical record was in Connecticut, and
- the hospital stay was associated with ICD-9-CM diagnosis code 079.6 (RSV), 466.11 (acute bronchiolitis due to RSV), or 480.1 (pneumonia due to RSV).

We designated epidemiological years as beginning in the first full week of July, in order to accommodate the timing of RSV peaks (which span two calendar years). Due to changes in diagnostic coding, we excluded data prior to July 1996; we similarly excluded data after June 2013 due to incompleteness of data for the 2013-14 epidemiological year. Using the weekly hospitalization data, we identified 9,701 RSV-associated hospitalizations among 300,693 total hospitalizations of patients <2 years old in Connecticut between July 1996 and June 2013 (inclusive).

### Spatial aggregation of RSV incidence

We analyzed RSV data aggregated either to the state level or into four geographic areas:

- One area for each of the counties with high population levels and high RSV case counts: Hartford, Fairfield, and New Haven (Figure 1).
- One discontiguous area comprised of all remaining counties in Connecticut: Tolland, Windham, Middlesex, Litchfield, and New London (“low-population counties”).

**Figure 1:**
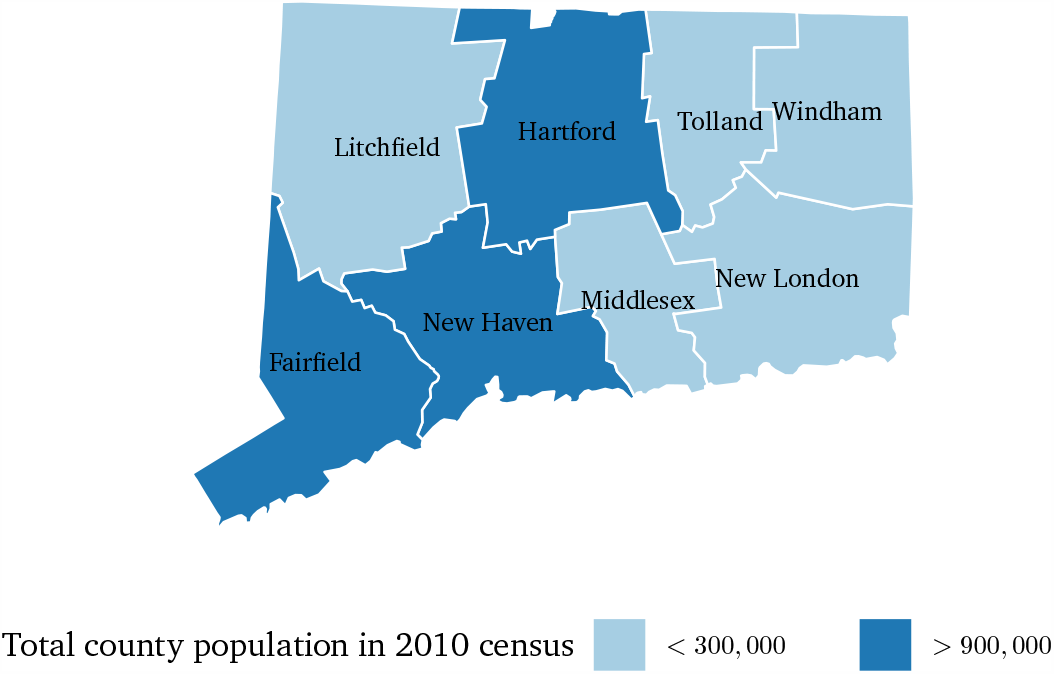
Subdivision of Connecticut into high-population and low-population counties

We chose county-level aggregation with an eye towards using this data to inform clinical practice guidelines, which are typically based on administrative areas for reasons of administrative and implementation simplicity. We chose to aggregate low-population counties into a single area because our analysis in those counties individually yielded results with low statistical power.

### Analysis approach

Our goal was to construct a model of annual RSV incidence that allowed us to estimate characteristics of RSV seasons, such as season onset, season offset, and preventable fraction of a prophylaxis regimen. To that end, we fit smooth curves through annual RSV hospitalization data and used resampling to obtain confidence intervals. This was performed using the pspline.inference package in R, which we developed for this purpose.^**methods**, 16^

We defined season onset as the time when cumulative incidence rises beyond 2.5% of the total cumulative incidence, and season offset as the time when cumulative incidence rises beyond 97.5% of the total cumulative incidence for an epidemiological year. We aggregated our data by surveillance week across all surveillance years, and estimated RSV season onset and offset for each county.

The choice of 2.5% and 97.5% incidence thresholds was based on our intent to use estimates of RSV season onset and offset to evaluate alternative prophylaxis regimens (as described below), while maintaining the 5-month dosing schedule (and therefore not increasing medication cost). Incidence cutoffs other than 2.5% and 97.5% were briefly investigated, but quickly ruled out because they produced prophylaxis regimens starting long prior to RSV season onset or ending long after RSV season offset, and therefore had very low preventable fractions.

### Evaluating alternative prophylaxis regimens

We compared the current AAP guidelines with six alternative prophylaxis regimens:

- **By current AAP national guidelines**: These guidelines recommend RSV prophylaxis with 5 monthly doses of palivizumab, starting in November, for all high-risk infants in their first year of life; in the second year of life, palivizumab is only recommended for some subgroups. For simplicity, we assumed that protection by palivizumab begins on November 15th.
- **By statewide onset**: prophylaxis administration begins at statewide median RSV season onset.
- **By statewide midpoint**: prophylaxis administration begins 12 weeks before the average of the statewide median season onset and offset.
- **By statewide offset**: prophylaxis administration begins 24 weeks before the statewide median RSV season offset.
- **By county-level onset**: prophylaxis administration begins at county-level median RSV season onset.
- **By county-level midpoint**: prophylaxis administration begins 12 weeks before the average of the county-level median RSV season onset and offset.
- **By county-level offset**: prophylaxis administration begins 24 weeks before the county-level median RSV season offset.

In all these regimens, prophylaxis consists of five monthly doses of palivizumab, and protection is assumed to last 24 weeks from the first dose. We rounded each alternative regimen to the nearest week to more closely reflect how prophylaxis guidelines might be implemented in practice (Supplemental Methods S1.1).

We defined the fraction of all RSV cases that is preventable by a prophylaxis regimen (“preventable fraction”) to be the ratio of

- the number of RSV-associated hospitalizations that occur in a given region (state or county) while the prophylaxis regimen offers protection to those who receive prophylaxis as scheduled, and
- the total number of RSV-associated hospitalizations that occur in the same region during a single epidemiologic year.

Given that high-risk infants are a small subgroup of the general population, we assumed that our model of RSV hospitalizations among all infants is an unbiased approximation of RSV hospitalizations among high-risk infants, and therefore that the preventable fraction of a prophylactic regimen is an unbiased estimate of the fraction of high-risk infants who benefit from prophylaxis.

### Additional analyses

In addition to the 1-week rounding of our alternative prophylaxis schedules, we also calculated schedules using 2- and 4-week rounding, to assess the impact of calendrical rounding on prophylaxis performance (Supplemental Methods S1.1).

To analyze the effects of year-to-year variation between RSV seasons, we performed a similar analysis, but with the prophylaxis start date in a given year determined by the median RSV season timing during the preceding three years, rather than all years (Supplemental Methods S1.2 and S1.3).

## 3 Results

### Timing of the RSV season varies within the state

Statewide, the median onset of the RSV season is at week 17.65 (95% CI: week 17.18 – 18.06), and offset is at week 41.71 (95% CI: week 41.35 – 42.13). The earliest season onset occurred in Fairfield county, with median at week 16.29 (95% CI: week 15.08 – 17.19). The latest season offset also occurrent in Fairfield county, with median at week 43.09 (95% CI: week 42.25 – 44.01). Meanwhile, the latest season onset occurred in the low-population counties, with median at week 20.07 (95% CI: week 17.18 – 18.06); the season offset in these counties was not different from the statewide median (Figure 2).

**Figure 2:**
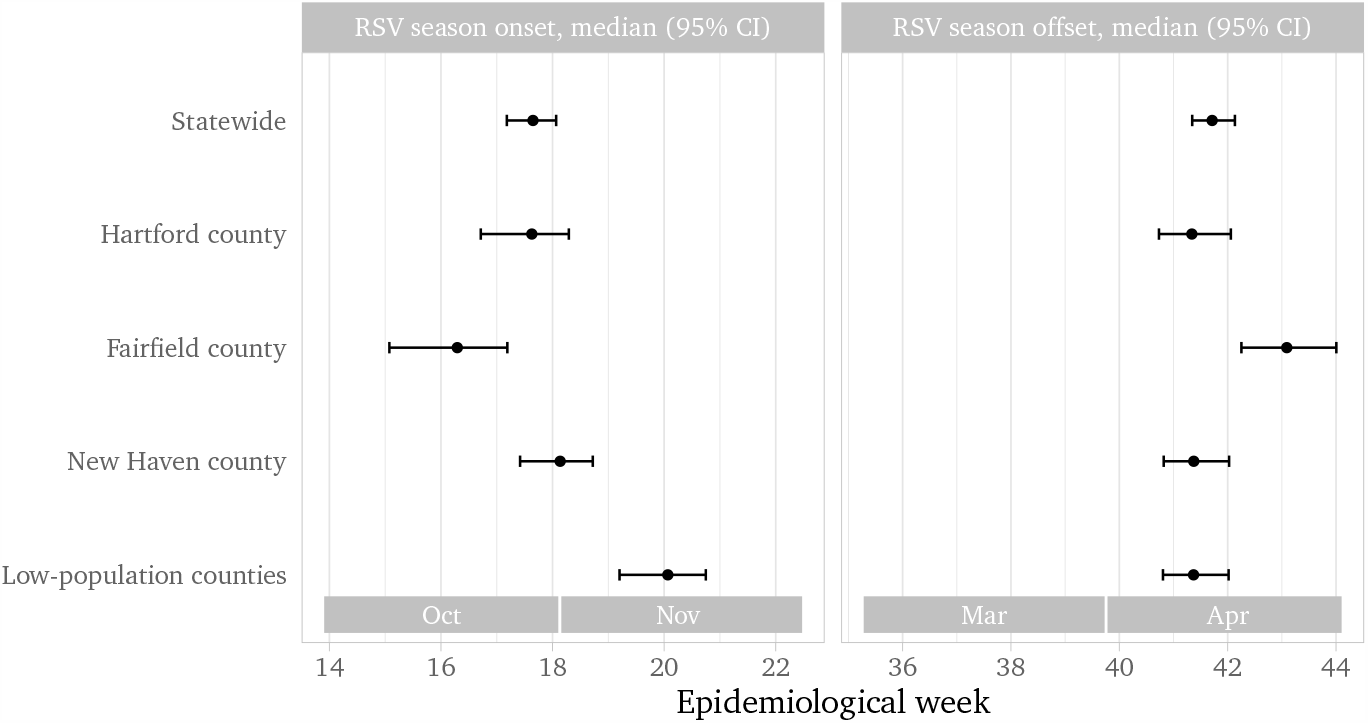
County-level variation in RSV season onset and offset in Connecticut

### The AAP guidelines perform best in low-population counties and worst in Fairfield county

Statewide, our model estimates that 94.07% of RSV cases (95% CI: 93.65 – 94.46%) occur while palivizumab offers protection when administered per the AAP guidelines. The AAP guidelines perform better in the low-population counties, where 96.61% cases (95% CI: 95.84 – 97.24%) occur during the prophylaxis interval, but worse in Fairfield county, where 91.89% cases (95% CI: 90.89 – 92.80%) occur during this time (Figure 3).

**Figure 3:**
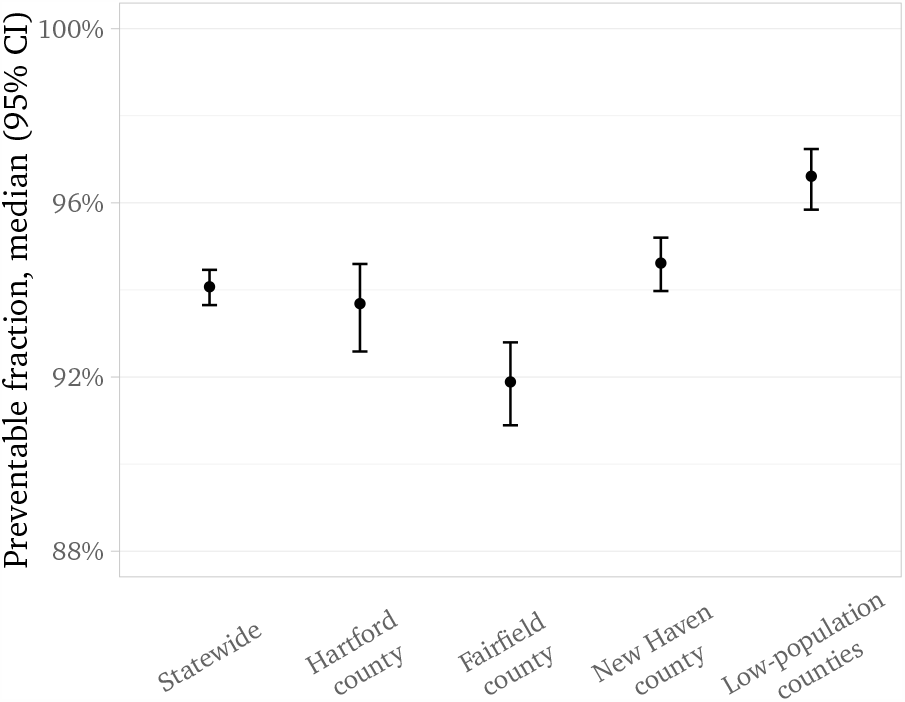
County-level variation in the preventable fraction of RSV hospitalizations according to AAP guidelines. The percentage of RSV hospitalizations occurring while protection by palivizumab — administered per the AAP guidelines — is active is plotted for each geographic region in Connecticut.

The county-to-county to variation in the protected fraction of the AAP-recommended prophylaxis is caused by temporal misalignment between the RSV season (delineated by median 2.5% onset and 97.5% offset in each county) and the timing of the AAP-recommended prophylaxis regimen (Figure 4). Fairfield county, owing to its early season onset, sees disproportionately more cases before prophylaxis administration begins; on the other hand, the low-population counties, owing to their late season onset and relatively shorter RSV seasons, see almost all of their RSV cases within the prophylaxis window.

**Figure 4:**
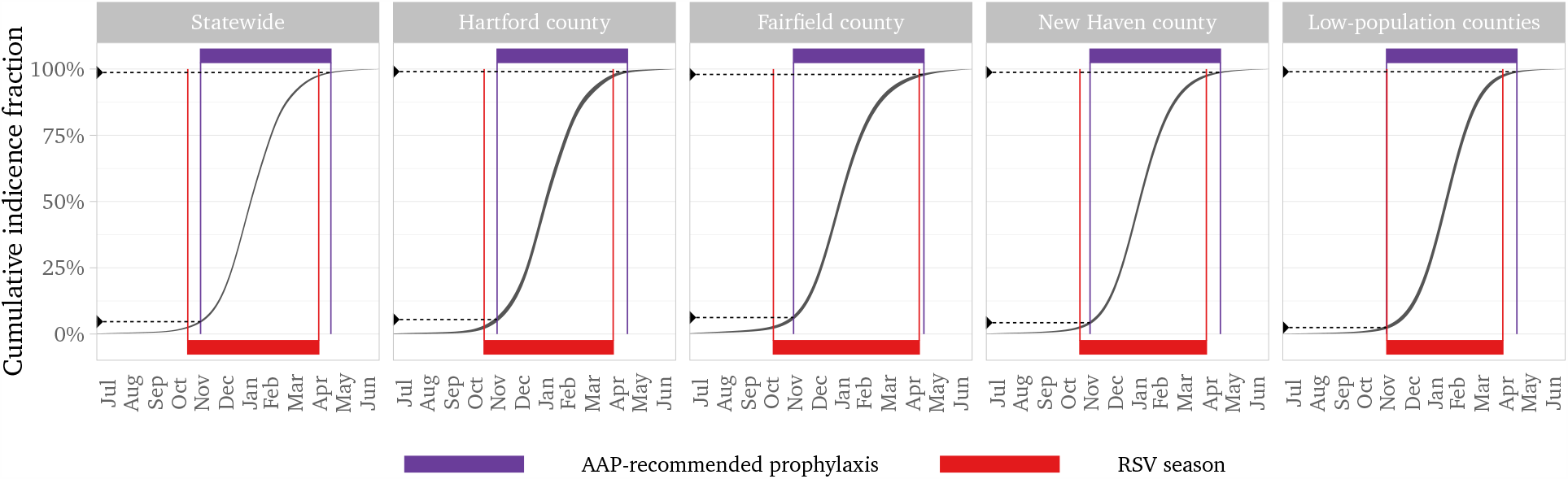
County-level variation in the relative timing of the RSV season in Connecticut and the AAP-recommended prophylaxis regimen. Comparison of the RSV season (red bar) and the prophylaxis window (purple bar) shows the misalignment between the two, notable particularly in Fairfield county.

### Spatially adjusted regimens are superior to the AAP recommendations

The six spatially-adjusted prophylaxis regimens yield results in all counties that are as good or better than the AAP guidelines. (Figure 5). Statewide, the AAP guidelines offer protection to 94.07% of cases (95% CI: 93.65 – 94.46%). The most favorable of our alternative regimens — based on statewide midseason — yields an increase in the preventable fraction to 95.07% (95% CI: 94.67 – 95.44%); other alternative regimens yield similar results. Most of the increase occurs in Hartford county, where the AAP guidelines protect 93.69% of cases (95% CI: 92.59 – 94.60%), whereas the most favorable of our alternative regimens — also based on statewide midseason — generates an increase to 95.23% (95% CI: 94.25 – 96.04%), with other alternative regimens being comparable. In every county, all alternative regimens are non-inferior to the AAP guidelines and non-superior to each other.

**Figure 5:**
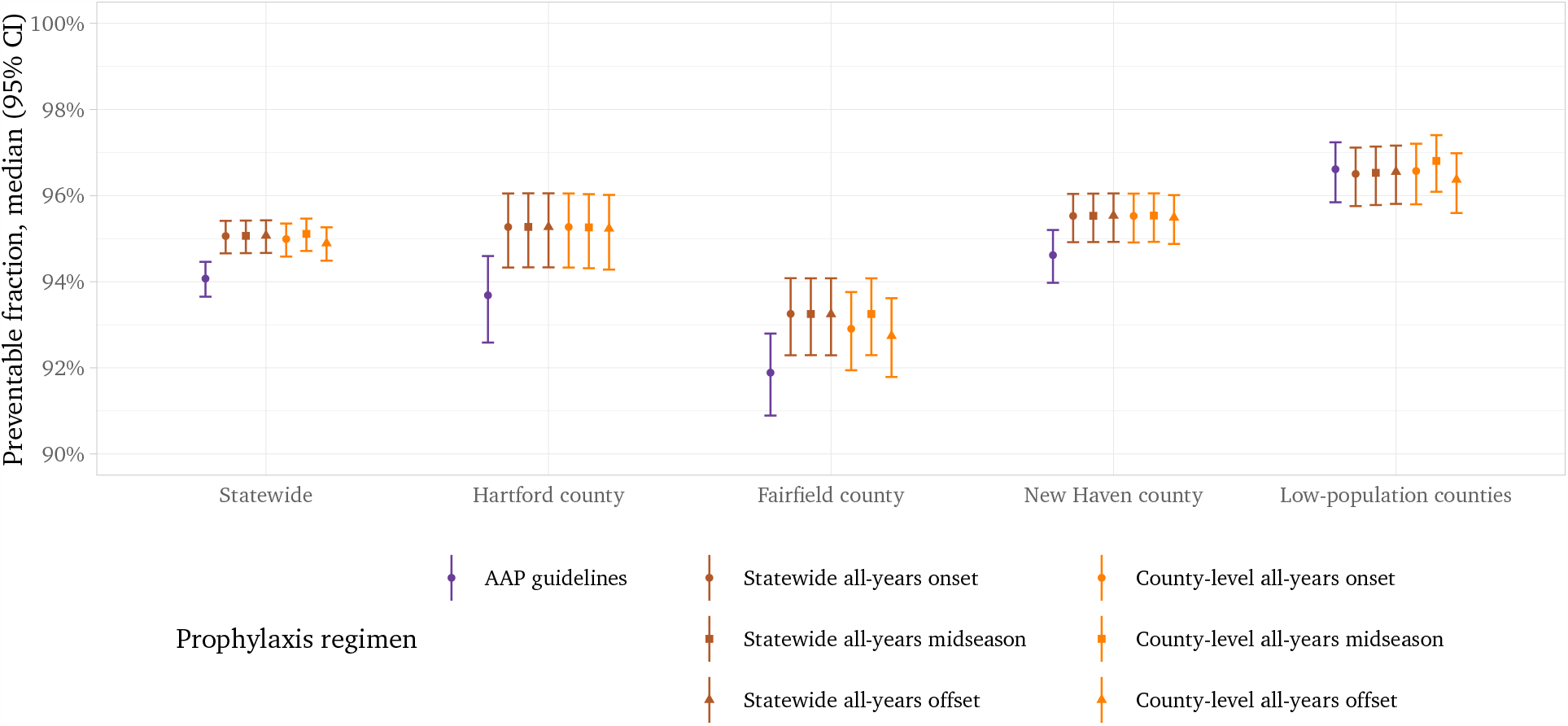
Comparison of six spatially adjusted RSV prophylaxis regimens to the AAP-recommended regimen. Spatially adjusted propxylaxis regimens are either non-inferior (for county-level analysis) or superior (for statewide analysis) to the AAP-recommended regimen. Preventable fraction: the fraction of all RSV-associated hospitalizations occurring while the prophylaxis regimen provides protection

### Additional results

We also applied 2-week and 4-week rounding to the start and end dates of our six alternative immunoprophylaxis regimens, and found that doing so diminished or erased the potential benefits (Supplemental Results S2.1). Finally, we analyzed long-term trends in the RSV season onset and offset and found that the season onset may be moving earlier (Supplemental Results S2.2), but that accounting for that drift did not yield a significant further increase in preventable RSV cases (Supplemental Results S2.3).

## 4 Discussion

Our analysis confirms the existence of county-level variation of RSV season timing in Connecticut (defined by RSV season onset and offset) observable at the county level. We have also shown a misalignment between the prophylaxis schedule recommended by the AAP and the timing of the RSV season throughout Connecticut, with most counties’ RSV season starting before protection from the first dose of prophylaxis is administered, and ending before protection from the last dose wanes. This misalignment is particularly notable in Fairfield County due to its early season onset.

Our effort to optimize timing of the prophylaxis regimen (without changing its duration and therefore its pharmaceutical cost) shows a potential ∼1% statewide decrease in RSV-associated hospitalizations among high-risk infants each year, primarily in counties with higher RSV burden (Fairfield, Hartford, and New Haven counties). The prophylaxis regimen with which we were able to attain the best improvement is the one in which prophylaxis in each county is timed relative to the midpoint of the RSV season in that county, which simultaneously reduces unprotected cases early in the season and ineffective prophylaxis late in the season. Nevertheless, all of the alternative regimens yielded similar results, with the timing of prophylaxis adjusted in each county or statewide by less than a whole month from the current AAP guidelines. Increasing complexity of practice guidelines would carry with it increased cost of implementation (such as training of clinicians and other healthcare workers); it is therefore not apparent from our research alone that the potential benefits would outweigh the costs.

Although we found a weak statewide trend in RSV season onset becoming earlier over time, further analysis did not yield any improvements to the preventable fraction of the prophylaxis regimen based on that trend. Since temporal variation in clinical guidelines would lead to a higher implementation burden than spatial variation, we see no reason to base RSV immunoprophylaxis clinical practice guidelines on temporal variation in RSV season timing.

The use of data aggregated by epidemiological weeks created a challenge for both our analysis and the interpretation of our results. In practice, clinical guidelines are typically expressed in terms of calendar weeks or months; as a result, our analysis contains errors of up to ±0.5 weeks when applied to the more natural calendar boundaries of clinical guidelines. Ideally, our analysis should be repeated with daily case counts aggregated into weeks and months that match week and month boundaries used in clinical guidelines.

Throughout, we assumed that RSV-associated hospitalizations are an unbiased estimate of RSV incidence in the general population. In doing so, we implicitly assumed that infectiousness and virulence of RSV are constant over time, for which we have no verification.

Our model of palivizumab immunoprophylaxis assumes a constant protection period with a hard start at administration and a hard stop at 24 weeks post-administration. The physiologic response to monoclonal antibody immunoprophylaxis is actually tapered on both ends. Similarly, we assumed the protection conferred by palivizumab is constant at 100% throughout the protection period; however, not only is breakthrough RSV illness known to occur in patients on palivizumab, but it is an indication for discontinuing the prophylaxis regimen.

In areas of Connecticut associated with a relatively high amount of interstate travel (such as urban centers near the state lines), our analysis — by only considering residents of Connecticut — likely underestimates RSV burden. Finally, towns that see substantial inter-county travel (for example, employment hubs near county lines) likely bias our county-level analysis.

In summary, our analysis reveals the potential for a ∼ 1% gain in preventable RSV cases in Connecticut when a locally-tailored two-week (rather than monthly) prophylaxis guideline is used in place of the established AAP guidelines. At an average of 571 RSV-associated hospitalizations per year in children under 2 years old in Connecticut, this corresponds to ∼5.71 hospitalizations prevented per year, and an unknown number of prevented cases not necessitating hospitalization. Practical aspects of this potential gain — such as implementation cost — require further investigation. Additionally, the method we developed for this analysis can be applied to county-level or state-level analysis of seasonal RSV patterns in other regions, some of which are likely to show a greater relative benefit (due to a greater degree of misalignment between the RSV season and the AAP guidelines’ prophylaxis window) as well as a greater absolute benefit (due to population size) than Connecticut.

## Data Availability

The data can be requested through the Connecticut Department of Public Health. The code is available at http://github.com/weinbergerlab/rsv-ct-optimization.git

## Supplements

### 1 Supplemental methods

#### S1.1 Accommodating implementation complexity

We performed three variants of our spatial analysis:

- **With weekly rounding**: prophylaxis start and end date rounded to the nearest calendar week,
- **With biweekly rounding**: prophylaxis start and end date rounded to the nearest two-week calendar period, with periods chosen to approximately align with November 15th, and
- **With monthly rounding**: prophylaxis start and end date rounded to the nearest four-week calendar periods, with periods chosen to approximately align with November 15th.

Alignment of rounding periods to November 15th was done to make our regimens more clearly comparable to the AAP-recommended regimen, whose (average) start date is also November 15th. This alignment of rounding periods was only approximate because we were working with data aggregated by epidemiological weeks, which aren’t aligned to calendar months.

#### S1.2 Temporal aggregation of RSV incidence

To analyze the effects of year-to-year variation between RSV seasons on the preventable fraction of prophylaxis regimens, we aggregated our data into (overlapping) periods of three consecutive years. We chose to use three-year periods in order to maintain a higher level of statistical power, as well as to limit the impact of short-term variation on our results. This gave us 16 three-year periods, with the 1998 period covering surveillance years 1996 through 1998, and the 2013 period covering 2011 through 2013.

We then estimated the onset and offset for each three-year period, using the same penalized spline generalized additive model as for the all-years analysis, producing — as before — statewide and county-level estimates of 2.5% onset and 97.5% offset. To evaluate year-to-year variation in RSV season onset and offset, we modeled median RSV three-year onset and offset against time using simple linear regression. We did not analyze any other long-term patterns, such as multi-year cycles.

#### S1.3 Evaluating alternative recent-years prophylaxis regimens

Finally, to assess the impact of any long-term trends in RSV season timing on prophylaxis, we evaluated three additional prophylaxis regimens. Here, each prophylaxis regimen is adjusted annually based on RSV data from the preceding three-year period:

- **By statewide recent-years onset**: prophylaxis administration in a given year begins at statewide median onset from the preceding three years.
- **By statewide recent-years midpoint**: prophylaxis administration begins 12 weeks before the average of the (statewide median) onset and offset from the preceding three years.
- **By statewide recent-years offset**: prophylaxis administration in a given year begins 24 weeks before the (statewide median) offset from the preceding three years.

We then compared those regimens to the one recommended by the AAP and to the six spatially adjusted alternatives (with no calendrical rounding) from our main analysis. As in the main analysis, these alternative regimens involved five monthly doses of palivizumab providing 24 weeks of protection.

### 2 Supplemental results

#### S2.1 County-level recommendations are most useful when rounded to the nearest 1- or 2-week period

After we applied weekly, biweekly, and monthly rounding to start and end dates of our six alternative immunoprophylaxis regimens, we found that longer rounding intervals diminished the benefit of tailoring the guidelines to local data (Supplemental figure S1). With weekly rounding, our regimens were non-inferior to the AAP recommendation, and produced an increase in preventable fraction from 94.07% (95% CI: 93.65 – 94.46%) under the AAP guidelines to 95.07% (95% CI: 94.69 – 95.44%) statewide. Biweekly rounding yielded regimens that were similarly non-inferior to the AAP recommendation, with an increase in the preventable fraction to 95.07% (95% CI: 94.67 – 95.43%) statewide. However, with monthly rounding, the gains almost completely vanished, with the preventable fraction not exceeding 94.53% (95% CI: 94.10 – 94.91%). Furthermore, all our regimens with monthly rounding were non-superior to the AAP-recommended regimen in all counties.

**Figure S1:**
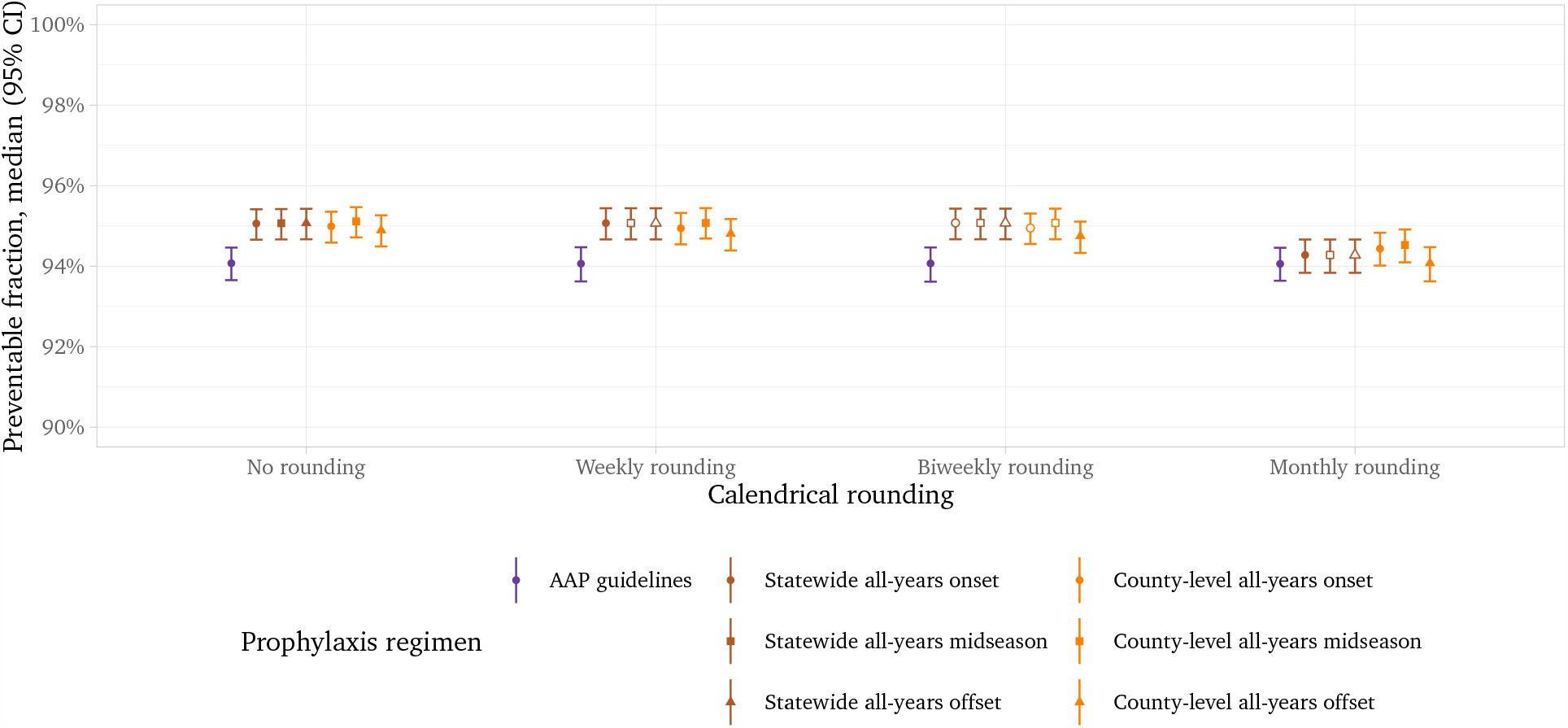
The effect of calendrical rounding of start and end of spatially adjusted RSV prophylaxis regimens on their performance. The preventable fraction (i.e. the fraction of all RSV hospitalizations occurring while the prophylaxis regimen offers protection) is plotted for each prophylaxis regimen and for each rounding interval. Our results show that longer rounding intervals diminish the benefits of alternative regimens over the AAP-recommended regimen. Hollow plot markers (○, □, and Δ) denote a prophylaxis strategy which, as a result of rounding, is identical to another, and therefore does not provide additional benefit.

#### S2.2 RSV season onset is slowly moving earlier

Our linear regression of median RSV season onset and offset over three-year periods indicates that season onset has slowly been drifting earlier (Supplemental figure S2), at a rate of −0.91 days/year (95% CI: −2.03 – 0.21 days/year). It is unclear whether this is a part of a larger pattern, such as a cycle spanning decades. Season offset, on the other hand, has not been drifting earlier or later (95% CI: −0.81 – 0.56 days/year).

**Figure S2:**
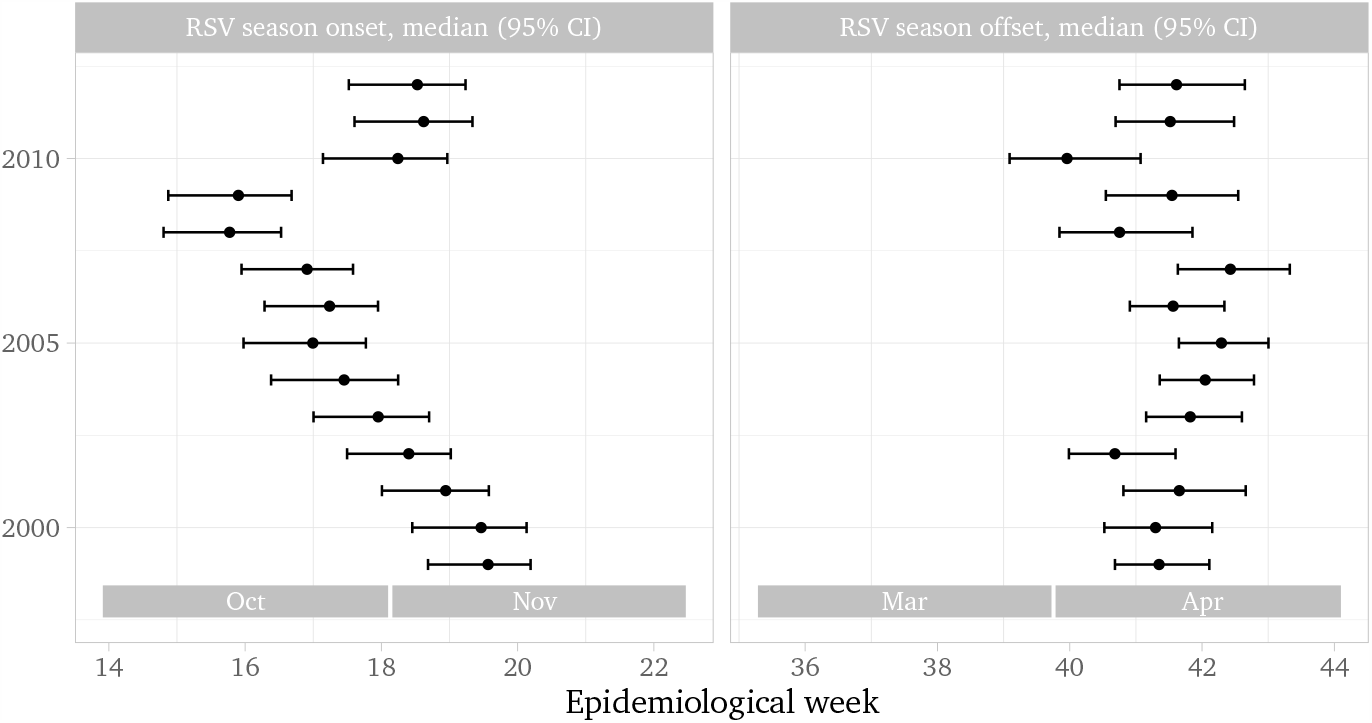
Annual variability in RSV season onset and offset in Connecticut. While season offset has been stable, season onset has shown a statistically significant drift toward an earliear RSV season.

#### S2.3 Temporally-adjusted prophylaxis regimens are non-superior to others

Compared to our county-level all-years analysis, the three regimens based on statewide recent-years analysis produced non-superior results everywhere and inferior results in the low-population counties (Supplemental figure S3). Due to the non-superiority of regimens based on statewide recent-years analysis, we did not evaluate regimens based on county-level recent-years data.

**Figure S3:**
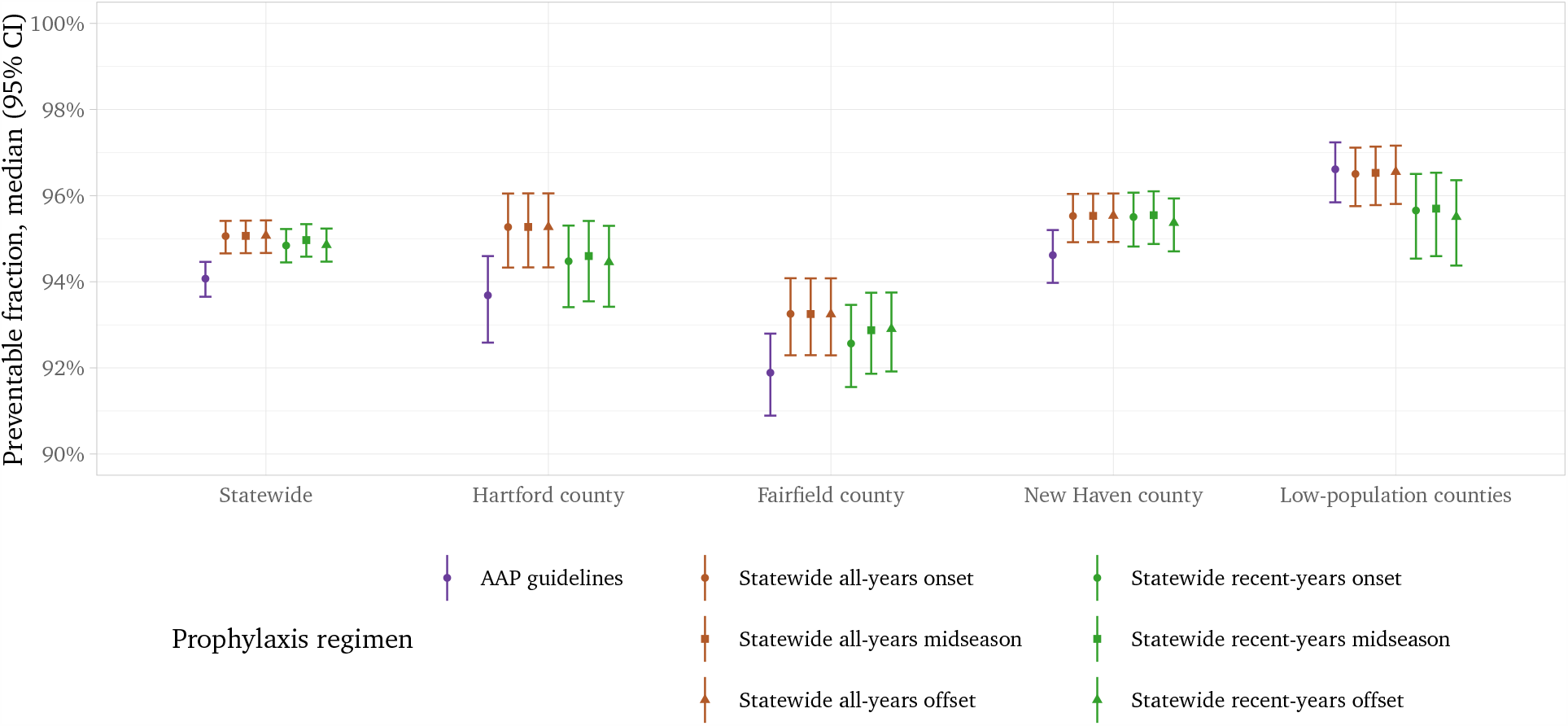
Comparison of prophylaxis regimens based on all-years data to those based on recent-years data. Recent-years analysis analysis adds no benefit over all-years analysis, for either statewide or county-level analysis. Preventable fraction: fraction of RSV hospitalizations occurring while the prophylaxis regimen offers protection

